# COVID-19 real world data infrastructure: A big data resource for study of the impact of COVID-19 in patient populations with immunocompromising conditions

**DOI:** 10.1101/2024.09.08.24313270

**Authors:** James M. Crawford, Lynne Penberthy, Ligia A. Pinto, Keri N. Althoff, Magdalene M. Assimon, Oren Cohen, Laura Gillim, Tracy L. Hammonds, Shilpa Kapur, Harvey W. Kaufman, David Kwasny, Jean W. Liew, William A. Meyer, Shannon L. Reynolds, Cheryl B. Schleicher, Suki Subbiah, Catherine Theruviparampil, Zachary S. Wallace, Jeremy L. Warner, Suhyeon Yoon, Yonah C. Ziemba

**Affiliations:** Northwell Health, Department of Pathology and Laboratory Medicine, New Hyde Park, NY; Division of Cancer Control and Population Sciences, National Cancer Institute, National Institutes of Health, Bethesda, MD; Frederick National Laboratory for Cancer Research, Frederick, MD; John Hopkins Bloomberg School of Public Health, Department of Epidemiology, Baltimore, MD; Aetion Inc., New York, NY; Laboratory Corporation of America, Burlington, NC; HealthVerity, Philadelphia, PA; Department of Pediatrics, Boston Children’s Hospital, Boston, MA (previously Quest Diagnostics, Secaucus, NJ); Boston University Chobanian & Avedisian School of Medicine, Section of Rheumatology, Boston, MA; Quest Diagnostics, Secaucus, NJ; LSU Health Sciences Center, Section of Hematology/Oncology, New Orleans, LA; Massachusetts General Hospital, Division of Rheumatology, Allergy, and Immunology, Boston, MA; Brown University, Department of Medicine, Providence, RI; Rhode Island Hospital, Providence, RI; Integrated Data Sciences Section, Research Technologies Branch, National Institute of Allergy and Infectious Diseases, National Institutes of Health, Bethesda, MD; Fortrea Inc., Durham, NC

**Keywords:** COVID-19, SEER, Cancer, Rheumatic Disease, Transplantation, Immunocompromised, CRWDi

## Abstract

**Background:** We created a United States-based real-world data resource to better understand the continued impact of the COVID-19 pandemic on immunocompromised patients, who are typically under-represented in prospective studies and clinical trials. **Methods:** The COVID-19 Real World Data infrastructure (CRWDi) was created by linking and harmonizing deidentified HealthVerity medical and pharmacy claims data from December 1, 2018 to December 31, 2023, with SARS-CoV-2 virologic and serologic laboratory data from major commercial laboratories and Northwell Health; COVID-19 vaccination data; and for patients with cancer, 2010 to 2021 National Cancer Institute Surveillance, Epidemiology, and End Results registry data. **Results:** The CRWDi dataset contains data on 5.2 million people. Four populations were included in the dataset: (1) patients with cancer (n=1,294,022); (2) patients with rheumatic conditions receiving pharmacotherapy (n=1,636,940); (3) non-cancer solid organ (n=249,797) and hematopoietic stem cell (n=30,172) transplant recipients; and (4) people from the general population including adults (≥18 years of age; n=1,790,162) and pediatric patients (<18 years of age; n=198,907).

**Conclusions:** We have created a complex real-world data system to address unanswered questions that have arisen during the COVID-19 pandemic. Further, by making the data broadly and freely available to academic researchers from the United States, the CRWDi real-world data system represents an important complement to existing consortia studies and clinical trials that have emerged during the healthcare crisis, and is readily reproducible for future purposing.

**Summary:** The COVID-19 Real World Data infrastructure dataset contains 5.2 million deidentified patient records, with focus on immunocompromising conditions, and is freely available to approved researchers to study the impact of coronavirus disease 2019 (COVID-19) on patient morbidities and outcomes.

## Introduction

Over the course of the COVID-19 pandemic, large-scale real-world datasets have enabled study of SARS-CoV-2 infection and outcomes within healthy and high-risk populations. A critical population of interest is immunocompromised patients, who either have diseases such as cancer or autoimmunity that impair their immune system or are receiving treatments that do so. These individuals have an increased risk for contracting infections, suffering poorer outcomes, and generating and spreading new SARS-CoV-2 variants, constituting a public health risk (1). These immunocompromised patients were generally excluded from participation in the initial COVID-19 vaccine immunogenicity and efficacy trials (2,3). Existing real-world datasets have left many questions unanswered pertaining to assessment and treatment of immunocompromised patients (4,5). These include understanding the chronology of SARS-CoV-2 infection and persistent infection or reinfection in such patients, the prognostic importance of the host serologic response to either natural infection or vaccination in predicting the severity of COVID-19 illness and post-acute outcomes, and the evolution in an immunocompromised patient of comorbid conditions and of health resource utilization. Thus, while clinicians must make decisions on how best to treat or immunize such patients, evidence-based guidelines remain high-level and do not provide detailed recommendations relevant to this patient population (6).

We sought to leverage real-world data across multiple data sources and organizations from the United States, to establish a combined, linked, deidentified infrastructure that would be available to the broader research community to begin to address many of the unanswered questions related to SARS-CoV-2 infection, focusing on higher risk immunocompromised populations. These questions include the variability in immune response to vaccination (7,8), the efficacy of vaccination against development of symptomatic disease (9–11), the efficacy of vaccination in preventing post-acute sequelae of COVID-19 illness (12,13,14), and the potential impact of SARS-CoV-2 infection on cancer recurrence and on the occurrence of new cancer, a question that has been raised from investigational studies (15). Addressing these questions may support the development of evidence-based guidelines for diagnostic evaluation and vaccination for patients across varying risk strata. Moreover, the establishment of CRWDi serves as proof-of-principle for making available a harmonized national data resource that can inform an evolving public health emergency such as a pandemic, particularly for patients who carry risk profiles that differ from the general population.

## Materials and Methods

### CRWDi Development and Oversight

HealthVerity (Philadelphia, PA) established CRWDi under the guidance of a subject matter expert working group having representation from the National Cancer Institute (NCI), Centers for Disease Control and Prevention (Atlanta, GA), Frederick National Laboratory (Frederick, MD), Laboratory Corporation of America (Burlington, NC), Quest Diagnostics (Secaucus, NJ), and academic health system-based individuals with established expertise in COVID-19 real-world data research. HealthVerity and Aetion, Inc. (New York, NY) also were members of the working group, in their capacities as data provider and analytic experts, respectively. Working group meetings occurred from February to December 2023, informed by preliminary analytic data provided regularly by HealthVerity. With continued engagement of a smaller executive group, CRWDi was completed in May 2024 (https://seer.cancer.gov/data-software/crwdi).

### Data Sources

The core data was from HealthVerity Marketplace (HVM; https://healthverity.com/), and included patient-level linked medical benefits and pharmacy benefits claims data from commercial health data sources as well as vaccination records from public health sources.

SARS-CoV-2 laboratory testing data was obtained from two major commercial laboratories (Laboratory Corporation of America (Labcorp) and Quest Diagnostics) and from Northwell Health (New Hyde Park, NY). For the commercial laboratories, testing data was based both on provision of clinically indicated testing for the United States, and on surveillance testing performed on remnant blood specimens under contractual arrangements with the CDC (18). Northwell Health laboratory data was based on provision of clinically indicated testing. For all sources, SARS-CoV-2 Nucleic Acid Amplication Test (NAAT) data was qualitative only (“detected” or “not-detected”) and did not include cycle threshold (Ct) values from the polymerase chain reaction on relevant platforms. The serologic SARS-CoV-2 anti-S test data was both qualitative (“detected” or “not-detected”) and semi-quantitative. Northwell also contributed SARS-CoV-2 Anti-N IgG test data, both qualitative and semi-quantitative. Laboratory test data from all three sources had accompanying Logical Observation Identifier Names and Codes (LOINC), and for the Anti-S and Anti-N semi-quantitative testing data, units, reference ranges and thresholds for detection, as per the assay platforms used by each laboratory.

To substantially enhance the robustness of information regarding patients with cancer, data from the NCI Surveillance, Epidemiology, and End Results (SEER) Registry was linked to existing patient records in the CRWDi. SEER includes 17 population-based cancer registries, representing approximately 48% of the US population (https://seer.cancer.gov/). This level of coverage provides for analytic validity and generalization with equitable selection of subjects (16). Cancer data from diagnosis years 2010-2021 were linked, including data on patient demographics, primary tumor site, tumor morphology, stage at diagnosis, first course of treatment, and follow-up vital status. CRWDi was thus provided with SEER information regarding cancer diagnosed both before (2010 to 2019) and during the first two calendar years of the pandemic (2020 and 2021), concordant with the 2-year lag time for complete case reporting to the SEER program (17). The inclusion of patients with incident cancer well prior to the pandemic permits the opportunity to study the associated risks of SARS-CoV-2 infection in cancer survivors, including those with recurrent disease.

### CRWDI Inclusion Criteria

The dataset was contractually targeted to be 5.2 million patients.

### General Inclusion Criteria

Patient inclusion criteria are provided in **Table 1**. First, the patients must have preexisting deidentified records in HVM for the time interval December 1, 2018 to December 31, 2023. The starting date of December 1, 2018 provides healthcare data information pre-dating COVID-19, thereby providing valuable longitudinal information on baseline health resource utilization. Second, although patients of all ages (in years) were to be included, those greater than 89 years of age would be coded as age 90 years, which was required for privacy purposes. Third, patients were included in CRWDi only if their records were informed for COVID-19 status by one or more of the following elements: (a) claims-based ICD-10 coding for COVID-19 or COVID-19 related conditions (the code for a COVID-19 diagnosis, U07.1, began on April 1, 2020 (19), with positive predictive value of greater than 90% for hospitalized patients, and 70% to 77% for outpatients (20–22)); (b) SARS-CoV-2 virologic and/or serologic testing data (regardless of negative or positive result); and/or (c) COVID-19 vaccination status. Fourth, patients need to be enrolled in a health plan with both Medical and Pharmacy benefits, with closed claims data available. For those patients meeting the enrollment criterion (see next), open claims data would be made available for analysis purposes. Fifth, patients needed to have been enrolled in a health plan for at least 180 days during the time interval December 1, 2018 to December 31. The 180 days enrollment was chosen to increase the likelihood of having robust longitudinal claims data.

**Table 1.** Inclusion Criteria for COVID-19 Real World Data infrastructure (CRWDi)

| *General Inclusion Criteria* |  |
| --- | --- |
| HVM enrollment dates | December 1, 2018 to December 31, 2023 |
| Age | 0 to 89 years; age >89 years coded as age 90 |
| COVID-19 status available (and/or): | Medical and/or Pharmacy Claims ICD-10 coding for COVID-19<br>Laboratory testing data: *<br>SARS-CoV-2 Nucleic Acid Amplification diagnostic testing<br>SARS-CoV-2 Serologic testing (total IgG and/or anti-S IgG)<br>COVID-19 vaccination CPT coding |
| Claims data | Medical and Pharmacy Closed Claims;<br>Open Claims data made available for analysis of patients<br>Meeting the $\geq 180$ days inclusion criterion for Closed Claims |
| Enrollment duration | $\geq 180$ days continuous health plan enrollment within the period 12/1/2018 to 12/31/2023, on the basis of Closed Claims |
| *Hierarchically Applied Criteria* |  |
| Cancer Patients | All HVM patients meeting above inclusion criteria, who overlapped with the NCI SEER Registry (inclusion dates January 1, 2010 to December 31, 2021) |
| Rheumatic Patients** | Remaining HVM patients with ICD10 coding for Rheumatic disease conditions, and with Closed Claims coding for active pharmaceutical treatment for Rheumatic disease |
| Transplant Patients*** | Remaining HVM patients with ICD10 coding for solid organ transplant or hematopoietic stem cell transplant |
| General Population† | Remaining HVM patients that had laboratory SARS-CoV-2 serologic testing data; pediatric ages $\leq 18$ years sampled to limit to the % pediatric population in the Cancer-Rheumatic-Transplant cohorts (10%). As the number of both General Population adult and pediatric patients with serologic testing data did not complete the planned total CRWDi population of 5.2 million, remaining General Population sampled to bring General Population cohort into accordance with the age and sex distribution of 2020 U.S. Census, again limiting pediatric contribution to 10%. |
| *Northwell Health Criteria* |  |
| COVID-19 laboratory data available for deidentified patients in the HVM database | Laboratory testing data:<br>SARS-CoV-2 Nucleic Acid Amplification diagnostic testing<br>SARS-CoV-2 Serologic testing (total IgG; Apr 2020 to Apr 2021)<br>SARS-CoV-2 Anti-S Serologic testing (IgG; Apr 2021 to Apr 2023)<br>SARS-CoV-2 Anti-N Serologic testing (IgG; Apr 2021 to Apr 2023) |

Enrollment criteria were applied sequentially. All de-identified patient records were drawn from HVM; data received from commercial laboratories, NCI SEER Registry, and Northwell Health was harmonized with HVM patient records.
\*As available from: Laboratory Corporation of America Holdings (Burlington, NC) and Quest Diagnostics Incorporated (Secaucus, NJ).
\*\*Mutually exclusive relative to Cancer patients; does not include rheumatic patients captured as part of cancer patient cohort.
\*\*\*Mutually exclusive relative to Cancer and Rheumatic patients; does not include transplant patients captured as part of cancer or rheumatic patient cohorts.
†Mutually exclusive relative to Cancer, Rheumatic, and Transplant patients
Abbreviations: HVM, HealthVerity Marketplace; ICD-10, International Classification of Diseases, 10<sup>th</sup> Revision; NCI, National Cancer Institute; SEER, Surveillance, Epidemiology, and End Results Program.

The remaining inclusion criteria were applied sequentially and hierarchically.

### Cancer cohort

The cancer cohort was defined by all SEER patients with cancer of any type, incident during calendar years 2010 to 2021, who linked with HVM records meeting general inclusion criteria.

### Rheumatic cohort

Drawn from HVM records meeting general inclusion criteria and after extraction of the Cancer cohort, the cohort of rheumatic patients had: (a) a rheumatic condition coded for in either medical and/or pharmacy claims; and (b) received treatment for a rheumatic condition at some time during the required 180-day enrollment period. Patients with rheumatologic conditions were identified through claims-based ICD-10-CM data as reported by D’Silva (23) and Qian (24). We imported all patients with rheumatic conditions other than rheumatoid arthritis or spondyloarthritis. As the latter two conditions were the most common rheumatic diseases, sampling of patients with these two conditions was required in order to limit the total number of rheumatic patients to 1.7 million. This last number was selected to be able to include 2.0 million general population patients in CRWDi.

### Transplant cohort

Drawn from HVM records meeting the general inclusion criteria and after extraction of the cancer and rheumatic cohorts, the cohort of transplant patients had ICD-10-CM and/or ICD-10-PCS coding for solid organ transplant or hematopoietic stem cell (HSC) transplant. All such patients were imported based on the premise that transplantation of any type would generate patient cohorts of interest.

### General population

The remaining HVM patients who met the general inclusion criteria were considered a general population, serving as a source for extraction of subcohorts appropriate for specific comparative research studies. The 3 cohorts previously selected hierarchically (cancer, rheumatic conditions, transplant) had an aggregate age distribution of approximately 90% adult (age 18 years or older) and 10% pediatric (age less than 18 years), which differs from the 22% pediatric population of the United States reported by the 2020 Census (25). To optimize the ability of researchers to identify general population subcohorts of age appropriate to their studies of immunocompromised patients, the general population selected for CRWDi was also targeted to be 90% adult and 10% pediatric patients. Since a key goal of creating CRWDi was to obtain real world evidence on the potential value of SARS-CoV-2 serologic testing, the first extraction of general population patients was those who had SARS-CoV-2 serologic test results. As this first extract did not fully meet the target general population cohort size of 2.0 million patients, a further extract was obtained of general population patients meeting the general inclusion criteria but who did not have SARS-CoV-2 serologic test results to bring this number to 2.0 million.

### Northwell Health data

Northwell Health was chosen as a contributing health system, both for having SARS-CoV-2 virologic testing from the start of COVID-19 pandemic in the greater New York metropolitan area (beginning March 7, 2020; 26) and for having activated serologic testing for COVID-19 on April 20, 2020 (27). The goal was for Northwell Health to supplement COVID-19 testing data obtained from the commercial laboratory sources. For Northwell patients with SARS-CoV-2 virologic and/or serologic testing data whose deidentified records were already present in HVM, the general inclusion criteria of: (a) 180 days continuous enrollment in benefits plan based on closed claim records; and (b) COVID-19 status informed by HealthVerity and/or commercial laboratory data, were not applied.

### Creation of the CRWDi dataset

As a data aggregation company, HealthVerity utilizes a highly reliable Privacy-Preserving Record Linkage (PPRL) solution to allow for data sharing while maintaining privacy (28,29; https://surveillance.cancer.gov/reports/TO-P2-PPRLS-Evaluation-Report.pdf). Deidentified tokens are generated for each patient at each data source (30); token matching then takes place exclusively at HealthVerity against the much larger encrypted and tokenized dataset maintained by HealthVerity. While patient identities are not released from the data owner, HealthVerity utilizes an independent third-party reviewer to ensure that all data is HIPAA compliant and has minimal risk of re-identifiability.

### Data categories for CRWDi

A summarized list of the data sources and the key data categories is shown in **Table 2**. From medical claims records, ICD-10-CM codes relating to acquired immunodeficiency syndrome and Human Immunodeficiency Virus were excluded for privacy concerns. ICD-10-CM coding for comorbid conditions, and in turn for determining the Charlson Comorbidity Index, is based on published work (31). Detailed treatment and dates of administration and/or prescription fill for cancer, rheumatologic disease, transplantation, and comorbid conditions, as well as specific therapies for COVID-19, are based on the National Drug Code (NDC) system from pharmacy claims and healthcare common procedure coding system (HCPCS)/CPT codes from medical claims data. For pharmacy data obtained from the SEER Registry, specific NDC and HCPCS codes for anti-neoplastic agents are based on the CanMed System (https://seer.cancer.gov/oncologytoolbox/canmed/ndconc/). Vaccine administration is derived from pharmacy claims and state level public health data provided by health agencies, providers and pharmacies.

**Table 2.**
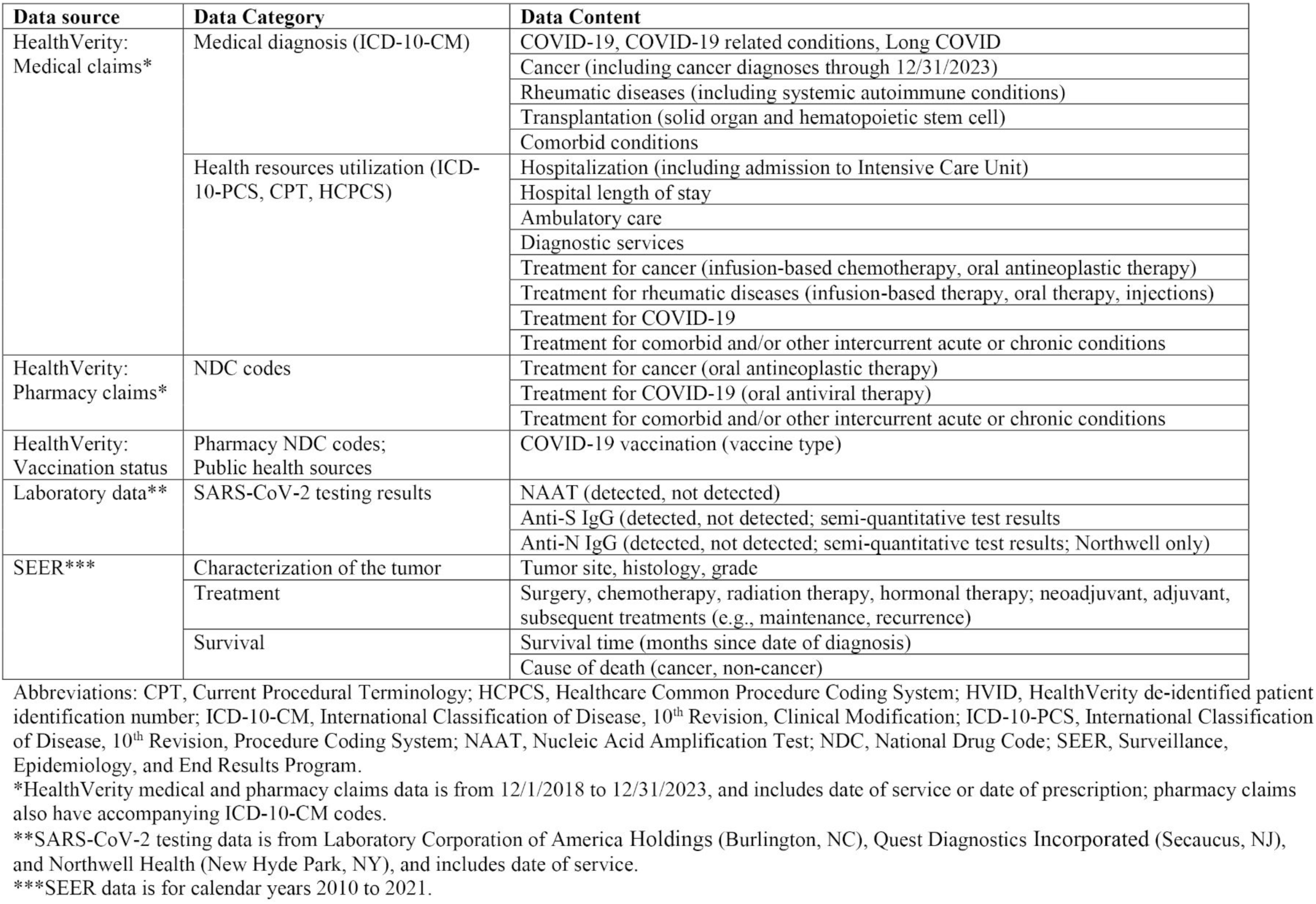
Data sources and data categories contributing to COVID-19 Real World Data infrastructure (CRWDi)

### Data Availability for Researchers

This data resource was created to support academic, non-commercial research projects in the United States. Submitted proposals are reviewed by the NCI for appropriateness of the proposal to the data resource (https://seer.cancer.gov/data-software/crwdi/). Upon approval, obtaining access to CRWDi requires both NCI-authorized access to the SEER Registry, and HealthVerity-authorized access to the cloud-based cohort discovery tool and analytic platform housing the CRWDi data.

The HVM analytics environment leverages the Databricks application (Databricks, Inc., San Francisco, CA) to provide researchers with a robust analytics layer. The CRWDi analytics platform capabilities include built-in support for multiple programming languages that are widely used in data analytics and machine learning, including Python, R, SQL, and Scala. Owing to privacy concerns, the cloud-based analytics precludes download of individual patient data but permits download of aggregated results such as tables and figures.

While the retrospective observational data are deidentified, the rich and longitudinal nature of the CRWDi data carries some risk of patient re-identifiability. Therefore, each research investigator requesting access to the CRWDi must obtain and show proof of institutional review board approval for a proposed study.

## Results

The CRWDi project was initiated in February 2023 and completed in May 2024. At the time of manuscript submission, research groups from six academic health systems in the United States have sought and obtained full permissions to utilize CRWDi.

The patient cohorts included in CRWDi are shown in **Figure 1**. The sum of the numbers in each grey box is exactly 5,200,000 unique patients. Owing to the hierarchical application of selection criteria, there is overlap between the first 3 cohorts. These overlapping relationships are shown in **Figure 2**. For researchers seeking to identify specific sub-cohorts of patients, the detailed diagnostic, procedure and pharmacy codes and laboratory data groups shown in Table 2 are available, as is the rich dataset from the SEER Registry.

**Figure 1.**
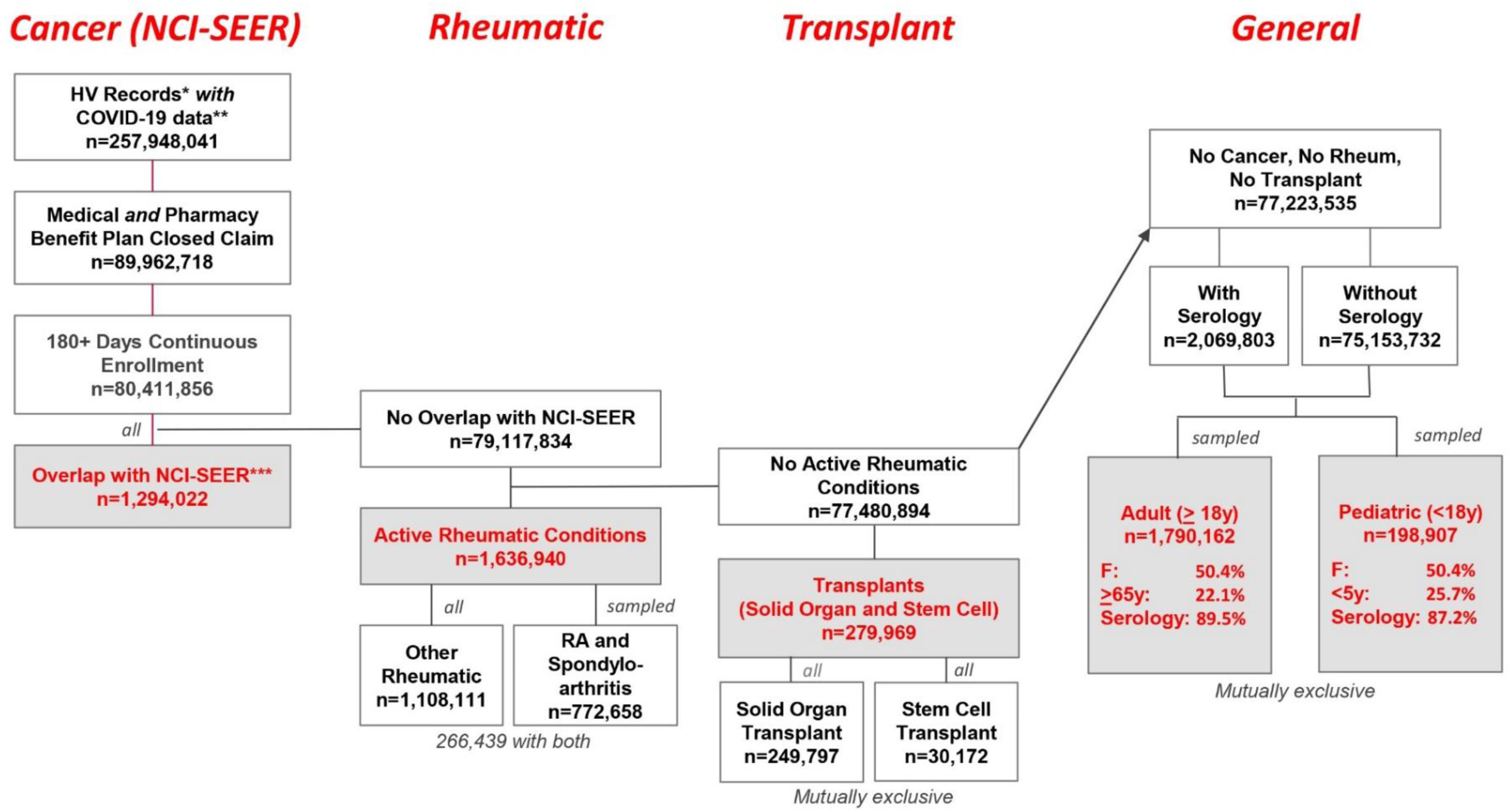
Waterfall chart of CRWDi cohort selection. Starting at the upper left, the sequential application of inclusion criteria presented in Table 1 yielded the population numbers shown in each box. The left column shows the extract for SEER Registry-informed cancer patients (grey box). The second column is the mutually exclusive remainder extracted for “active” rheumatic conditions (on medication during the minimum 180 day enrollment period). Owing to their large numbers, patients having active rheumatoid arthritis or spondyloarthritis were sampled for inclusion in CRWDi; all patients with other rheumatic conditions were included. Since ICD-10 coding might include patients in each subgroup, the actual number of unique rhematic patients is as given in the grey box; the number of patients having diagnostic codes both for other rheumatic conditions and for rheumatoid arthritis or spondyloarthritis is shown at bottom. The third column is the mutually exclusive remainder extracted for solid organ or hematopoietic stem cell transplantation; there was no overlap between the transplant subgroups. The fourth column is the mutually exclusive remainder, constituting a general population. The sampling of this population to obtain the numbers of unique adult and pediatric patients given in the grey boxes is given in the Methods. *HealthVerity Marketplace claims from 12/1/2018 to 12/31/2023. **HealthVerity Marketplace ICD-10 coding for COVID-19; and/or commercial laboratory testing data on SARS-CoV-2 Nucleic Acid Amplification Test virologic and/or SARS-CoV-2 serologic testing; and/or HealthVerity Marketplace coding for COVID-19 vaccination. ***NCI-SEER Registry data for calendar years 2010 to 2021. Abbreviations: HV, HealthVerity; NCI-SEER, National Cancer Institute SEER Registry; ICD-10, International Classification of Disease, 10^th^ Revision; RA, Rheumatoid arthritis; Rheum, Rheumatic.

**Figure 2.**
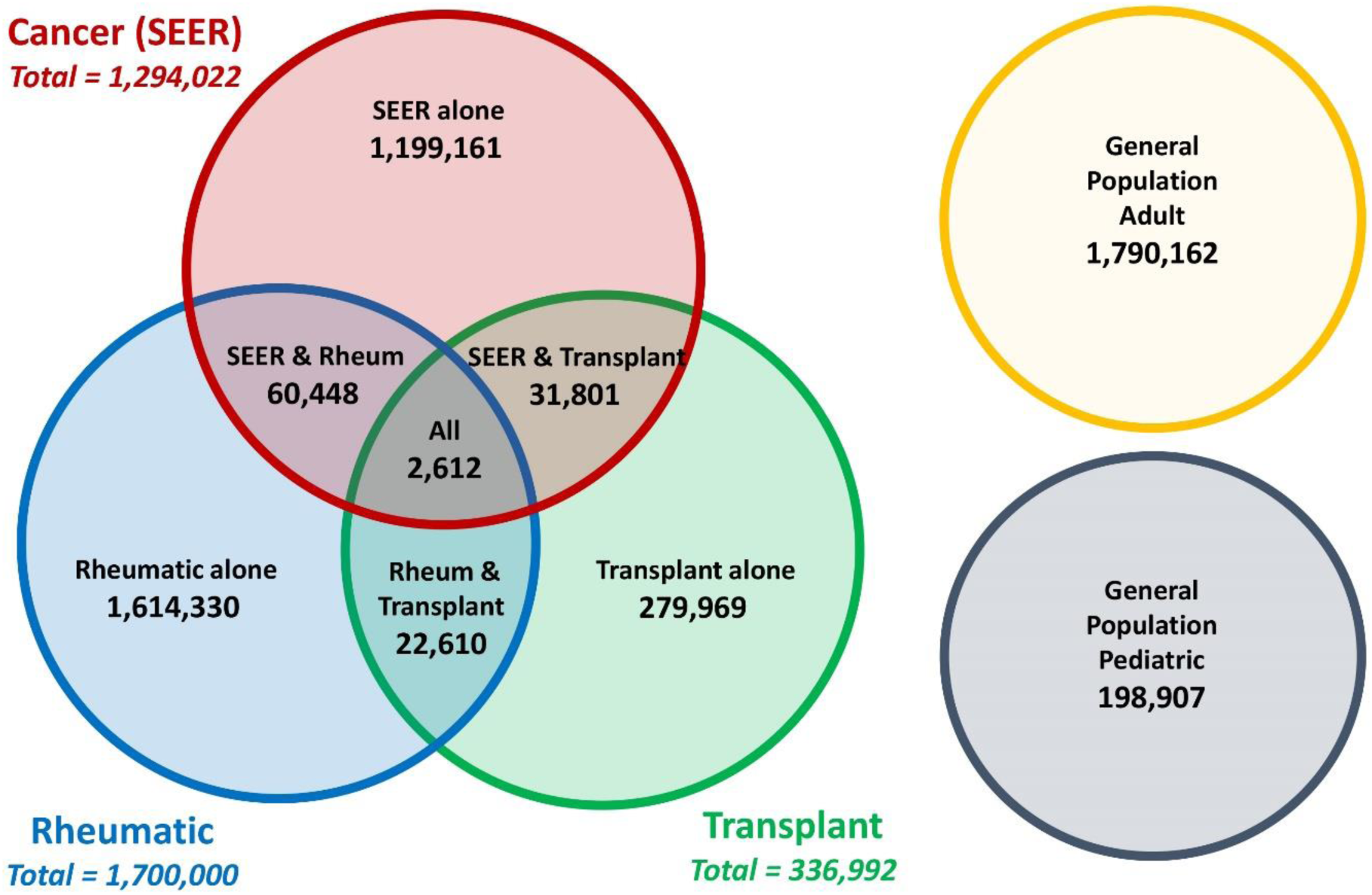
Venn diagram of CRWDi cohorts. Following the hierarchical cohort selection process shown in Figure 1, the final patient distributions between the three cohorts of immunocompromised patients are shown, to show the overlap of patients with more than one condition. The mutually exclusive adult and pediatric population cohorts also are shown. The circles are not to scale. The total number of deidentified patients totals exactly 5,200,000.

Since the SEER Registry represents approximately half of cancer patients in the United States, it is likely that some rheumatic and transplant patients may be patients with cancer who are not in the SEER Registry. Further, given that the SEER Registry was reconciled through December 31, 2021, it is possible that patients receiving an incident diagnosis of cancer between January 1, 2022 and December 31, 2023 would be identified only through the HVM dataset, and so would also potentially be included in the non-SEER patient cohorts shown in Figure 2. Such considerations would become evident during the analytic phase of any research project.

Separate from the CRWDi final cohorts shown in Figure 2, 336,545 unique Northwell patients were linked. Notably, these Northwell laboratory data supplemented 40,654 patient records already present in the 5.2 million CRWDi patient population shown Figure 2. However, an additional 295,891 unique patients were added to the CRWDi data resource based on Northwell laboratory data, who would not otherwise have been identified in HVM but now also are available for study.

## Discussion

The CRWDi infrastructure was developed by consolidation and linkage of multiple real-world data sources of patients in the United States, and is now available to the national research community (https://seer.cancer.gov/data-software/crwdi). Collectively, CRWDi constitutes a non-random sample of real-world data informed by COVID-19 status, on a substantive population of immunocompromised patients, with a comparator general population. The fact that the general inclusion criteria were for deidentified patients who had COVID-19 related data, including negative as well as positive laboratory test results for COVID-19 (virologic and/or serologic), means that research studies will be informed by patients who did experience SARS-CoV-2 infection, in comparison to those that did not.

Having been established, this data resource also can be updated. Further, by modifying the inclusion criteria, the CRWDi approach to dataset creation also can be replicated relatively quickly in the event of a different health crisis or to address other important clinical or public health questions. Importantly, the use of PPRL for the consolidation of the multiple data sources represents a relatively new but effective method to bring together data components that only together can answer questions not possible using a single data source, however large (30).

While real-world data cannot replace clinical trials, the ability to leverage real-world datasets allows investigators and governmental agencies to focus on important subpopulations who are unlikely to be eligible for clinical trials. CRWDi may also be useful to validate observations obtained from other large, national consortia, particularly those based solely on electronic health records. A strong advantage of CRWDi is identification through administrative claims of longitudinal data on patients who have received care from multiple health systems, and who have obtained and filled pharmacy prescriptions in the ambulatory setting. Indeed, information on ambulatory pharmaceutical treatment for both COVID-19 and co-existent conditions such as cancer or autoimmune disease provides a powerful enhancement for understanding the impact of COVID-19 in these higher risk patients (32). Although the claims-based foundation of CRWDi omits patients who do not have health insurance, this dataset is otherwise drawn from the entire U.S. population, and so should yield generalizable findings.

A limitation of the NCI SEER Registry is both that the data are through December 31, 2021 only, and that SEER data may be incomplete with regards to subsequent courses of therapy, disease progression, or recurrence (17). Linkage of SEER data to the claims data of HealthVerity provides access to extensive information about the longitudinal clinical course of cancer patients through December 31, 2023, thus substantively extending the chronologic reach of the SEER Registry or the SEER-Medicare database (17).

The integration of novel data sources such as the SEER Registry data and Northwell data supplements the existing HVM claims and commercial laboratory data in important ways. The SEER data represent carefully adjudicated information on incident cancer cases that are followed longitudinally, information that is not available through administrative claims data. In turn, by providing a large sample of serologic test results (n=376,259), the Northwell Health data provide critical opportunity for validation of the potential value of COVID-19 serologic testing. The fact that Northwell Health data identified an additional n=295,891 patients whose COVID-19 related laboratory data could be linked to the HVM demonstrates the potential importance of including data from health systems in development of a harmonized national data infrastructure for study of public health emergencies.

There are certain limitations of CRWDi. First, we required that a patient have continuous benefits enrollment for at least 180 days during the study period of December 1, 2018 to December 31, 2023 to assure comprehensive capture of information during the covered period. There is the potential of selection bias against patients who contracted COVID-19 and succumbed in less than 180 days. However, preliminary analysis of the CRWDi dataset shows that most patients with a history of cancer from the SEER Registry were enrolled in benefit plans for much longer periods (median enrollment was of duration >1500 days, data not shown). This suggests that benefits enrollment for immunocompromised patients is more likely to have predated an index date for SARS-CoV-2 infection, and hence accelerated severe courses of COVID-19 would be captured. Second, while one measure of diagnosis of infection with SARS-CoV-2 included presence of a positive NAAT, with the advent of home antigen testing a substantial portion of patients with SARS-CoV-2 infection who did not seek medical care may be missed, raising the possibility of bias towards null outcomes (33). However, since the scientific focus for CRWDi is to assess risk for severe disease and/or post-COVID-19 sequelae, it is likely that the CWRDi will have a high capture rate for this important subset of the SARS-CoV-2 infected population. Third, CRWDi relies on medical coding, e.g., ICD-10, NPC, and CPT, and reporting of cancer diagnoses (SEER). The limitations of administrative coding data are well-documented, with regards to potential diagnostic or procedural coding inaccuracies (34–36). The capture of health resource utilization data as well as diagnostic and treatment coding data should help overcome this potential limitation, by informing researchers on the health care needs of patients separate from the diagnoses those patients might be assigned.

There are other infrastructures that represent important data sources to study SARS-CoV-2 (37–42); these sources represent complimentary but distinct data consolidation efforts. Such data resources also are subject to the potential limitations of accuracy and reliability in administrative coding of health care encounters. The electronic health record-based consortia and the federal Sentinel program represent real-world data that provide a different perspective on the COVID-19 pandemic than the CRWDi, and thus these multiple systems are complementary to one another.

## Conclusion

The CRWDi infrastructure, consisting of COVID-19 related data based on healthcare claims and other data from United States patients, achieved the several key objectives for which the system was created. First, it demonstrated that a complex harmonized real-world data system can be created to address ongoing and new questions arising during a pandemic, that are not otherwise addressable by established data consortia. Second, CRWDi represents the ability to study important population subgroups that are typically under-represented in prospective cohort studies and clinical trials, but for whom important clinical questions remain unanswered. Further, the data have been made broadly freely available to academic researchers through an NCI-based proposal review-and-approval process. In summary, the CRWDi real-world data system represents an important complement to existing consortia studies and clinical trials that emerged during healthcare crisis, can be updated in continuation of its current purpose, and is reproducible for future purposing.

## Notes

## Acknowledgements

The authors thank these individuals who also contributed to CRWDi planning discussions: Tegan Boehmer, Commander, Adi Gundlapalli, Ph.D., Matthew Ritchie, Ph.D., of the Centers for Disease Control and Prevention, Atlanta, GA. These individuals from HealthVerity, Inc. also are thanked for their contributions: Rick Edwards, Christopher Williams, and Sean Thompson.

## Abbreviations

CDC: Centers for Disease Control and Prevention
CPT: Current Procedural Terminology
CRWDi: COVID-19 Real World Data infrastructure
HCPCS: Healthcare Common Procedure Coding System
HSC: Hematopoietic stem cell
HVID: HealthVerity de-identified patient identification number
HVM: HealthVerity Marketplace
ICD-10-CM: International Classification of Disease, 10^th^ Revision, Clinical Modification
ICD-10-PCS: International Classification of Disease, 10^th^ Revision, Procedure Coding System
LOINC: Logical Observation Identifier Names and Codes
NAAT: Nucleic Acid Amplification Test
NCI: National Cancer Institute
NDC: National Drug Code
PPRL: Privacy preserving record linkages
SEER: Surveillance, Epidemiology, and End Results program.

